# Right Thoracotomy with Central Cannulation for Valve Surgery: 10 Years of Experience

**DOI:** 10.1101/2024.03.30.24304889

**Authors:** Anil Sharma, Sunil Dixit, Dr.Mohit Sharma, Dr.Sourabh Mittal, Dr.Apurva Shah, Shefali Goyal

## Abstract

**Background:** The aim of this study is to report the early outcomes of valvular heart surgeries performed via the right thoracotomy approach. While thoracotomy with femoro-femoral bypass is an established method for minimally invasive open-heart surgeries, thoracotomy with conventional cannulation is still being explored. In our center, we conducted 958 valvular heart surgery cases using the right anterolateral thoracotomy approach with central cannulation and data were analyzed.

**Methods:** This is a retrospective observational study based on prospectively collected data from patients who underwent valvular heart surgery at our center spanning from April 2013 to April 2023. The data encompass demographics, procedures, operative techniques, post-operative morbidity, mortality, and a 1-month follow-up.

**Results:** Our study revealed no procedure-related mortality. No patient required conversion to median sternotomy. Smooth cannulation and satisfactory exposure were achieved in all patients. The study encompassed a wide age range, from 14 to 68 years, with 618 female patients (64.5%) and 340 male patients (35.5%).The average cross-clamp time ranged from 38 to 90 minutes, the duration of cardio-pulmonary bypass ranged from 45 to 105 minutes, post-operative extubation ranged from 3 to 8 hours, the average drain volume ranged from 100 to 350 ml, and the incision size ranged from 5 to 7 cm.

**Conclusions:** Our data demonstrate that conventional cannulation via the right antero-lateral thoracotomy approach for valvular heart disease is a viable alternative to reduce the side effects associated with sternotomy and femoral cannulation. This procedure is safe, reproducible, and provides the same level of treatment quality.

## INTRODUCTION

Traditionally, median sternotomy has served as the gold standard approach for open heart surgeries in cardiac surgery, consistently yielding excellent outcomes [1]. Over time, there has been a growing interest in developing minimally invasive techniques to enhance cosmetic results and improve patient comfort in the realm of adult cardiac surgery. These approaches include ministernotomy [2, 3], a transxiphoid approach without sternotomy, anterolateral thoracotomy [4, 5], and posterolateral thoracotomy [6], all designed to achieve optimal repair outcomes while minimizing the cosmetic impact.

As the clinical outcomes of cardiac surgery continue to advance, often achieving near-zero perioperative mortality rates, the aesthetic considerations associated with these procedures have gained prominence. Beyond cosmetic benefits, there is an expectation that minimally invasive approaches may offer advantages such as reduced postoperative pain, decreased bleeding, enhanced respiratory function, shorter hospital stays, and ultimately reduced overall costs. However, the extent to which these minimally invasive techniques genuinely deliver on these promises remains a topic of debate in the medical community.

In this retrospective study, we aim to review our 10 years of experience with a specific minimally invasive approach, involving right anterolateral thoracotomy and conventional central cannulation, in valvular open heart surgeries in a cohort of 958 patients. It is worth noting that anterolateral thoracotomy with central cannulation has been adopted at select centers worldwide for adult valvular open-heart surgeries [7,8].

## METHODS

This is a retrospective observational study of prospectively collected data of patients underwent open heart surgery for valvular heart disease in Cardio Vascular and Thoracic Surgery department at our tertiary level institute from April 2013 to April 2023. Intraoperative events and operative outcomes, monitored over time, were assessed using contingency table analysis. Mechanical and bio-prosthetic valve were used according to our institutional protocol. To gather additional insights, each patient was requested to complete a pre-structured subjective questionnaire during a 1-month follow-up. This questionnaire focused on their perception of surgical scarring and the timeline for their return to routine activities. Furthermore, with the patients’ consent, photographs were taken to complement the subjective assessments.

### Inclusion criteria

1. Patients with rheumatic heart disease (Mitral, Aortic, Tricuspid and Double valve)
2. Severely calcific mitral stenosis
3. Severe mitral regurgitation
4. Mitral valve disease with LA/LAA clot
5. Isolated tricuspid valve disease
6. Calcific aortic valve disease
7. Aortic valve regurgitation
8. Double valve -mitral and aortic
9. Double valve-mitral and tricuspid
10. Age more than 14 years less than 70 years

### Exclusion criteria

1. Valvular hear disease due to Congenital cause
2. Valvular heart disease due to Ischaemic cause
3. Ejection fraction < 30%
4. History of previous thoracic surgery
5. Associated lung disease
6. Active endocarditis with vegetation
7. Re-do procedures,
8. Extreme age (<14 and >80 years)
9. Emergency and atherosclerotic aorta
10. All contra-indications of valvular heart surgery

All patients had a transthoracic echo at the time of discharge. The main outcomes investigated were early mortality, perioperative complications, recurrence and need of re-operation. Early mortality was defined as any death occurring within 30 days of operation or before discharge from the hospital.

## Operative Procedure

### Anesthesia

We used the standard protocol of anesthesia, i.e., as for conventional valvular heart surgery under general anesthesia. The patients were intubated with a single lumen endotracheal tube.

### Positioning the patient

The patient was positioned in a supine posture, with a sandbag strategically positioned beneath the right scapula. This placement facilitated a slight elevation of the patient’s right chest, typically within a range of 30 to 40 degrees, as required by the surgeon to optimize the surgical field exposure (see Fig. 1). The patient’s right arm was gently positioned alongside the body. Additionally, the right groin area was prepared and appropriately draped, ensuring readiness for potential emergency scenarios.

**Fig 1.** a. Patient position b. Incision c. Cannulation

### Right Anterolateral Thoracotomy (RAT)

The surgical procedure began with a 5–9 cm sub-mammary incision, initiated in the right sub-mammary fold, positioned approximately 3 to 4 cm away from the lateral border of the sternum (refer to Figs. 1). The breast tissue was carefully mobilized, and access to the right thoracic cavity was achieved through either the third or fourth intercostal space. The choice of the intercostal space depended on the specific valve being addressed, with the third space preferred for mitral procedures, the second space for aortic procedures, and the second space for double valve (mitral and aortic) interventions. The selection of the intercostal space was determined through pre-operative chest X-ray assessment to optimize surgical field exposure.

To prepare for cardiopulmonary bypass, a single venous cannula was initially introduced, allowing for the decompression of the heart to create additional space (Fig-1). Subsequently, a second venous cannula was introduced to establish full bypass flows. A chest retractor was carefully positioned between the intercostal spaces and opened gradually and gently to safeguard the ribs. To expose the pericardial sac, the right lung was gently compressed with a moist sponge. The pericardial sac was accessed through a 3– 4 cm incision located anterior and parallel to the phrenic nerve, extending from the diaphragm to the aortic reflection. Multiple stay sutures were strategically placed to elevate the heart into the operative field.

The critical element of this procedure lies in achieving optimal surgical exposure. All operations were performed using standard cardiopulmonary bypass through central cannulation, maintaining a state of moderate hypothermia. Aortic and venous cannulation followed the same principles as those applied in standard sternotomy. The utilization of long snares (as depicted in Fig. 1) was instrumental in ensuring that the snares and associated artery forceps remained clear of the operative field, thereby preserving an unobstructed view. Double purse string sutures were employed for both aortic and cardioplegia cannulae, further enhancing surgical exposure.

### Elaborated surgical steps-

1. Preoperatively, the choice of the thoracotomy space (typically the third or fourth intercostal space for right anterolateral thoracotomy for mitral and the second intercostal space for aortic valve replacement and double valve replacement) was determined based on chest X-ray assessment.
2. Following the thoracotomy, the pericardium was sequentially hitched using silk stay sutures, a step-by-step process. Traction was applied to these stay sutures, directed outward and downward, thereby bringing the cardiac surface into the surgeon’s view (see Fig. 1).
3. Aortic cannulation presented a particular challenge. We employed the Medtronic arterial cannula (Flexible Arch Arterial Cannula, Manufacturer: Medtronic, Inc., Minneapolis, USA) and the Edward Life Sciences venous cannula (Thin-Flex Single Stage Venous Drainage Cannula, Manufacturer: Edward Life Sciences LLC, USA).
4. For aortic cannulation, we delicately grasped the epiaortic tissues using right-angled artery forceps to provide gentle traction, while another artery forceps secured the tip of the aortic cannula, ensuring a smooth insertion.
5. In cases where venous cannulation posed difficulties, particularly in large right atrium cases, we initiated bypass with a single cannula first to create ample space for unimpeded access to the superior vena cava and inferior vena cava.
6. Aortic cross-clamping was performed using either a Chitwood clamp or a conventional clamp, both accessed from the same surgical incision. The Chitwood clamp was secured with a silk stay suture to prevent any interference by the clamp in the surgical field.
7. We employed antegrade del Nido cardioplegia for myocardial preservation. The cardioplegia cannula was the same as used in conventional surgery, supplied by Edward Life Sciences/Medtronic.
8. The left atrium was opened in a parallel manner to the atrioventricular groove, and the aorta was opened horizontally and parallel to the annulus.
9. De-airing of the heart was conducted through the left atrium vent and aortic root vent before gradually weaning off bypass.
10. Single ventricular and single atrial pacing wires were placed for atrioventricular pacing. The pericardium was closed above the aorta and partially above the right atrium, with the remaining pericardium left open to facilitate drainage into the thoracic cavity. A single chest drain was inserted to manage drainage from both the thoracic and pericardial cavities.

### Statistical analysis

continuous data were expressed as mean ± standard deviation. Categorical data were expressed as percentage. All statistical analyses were performed with SPS 22.0 (SPSS, Inc., Chicago, IL, USA).

## RESULTS

In this study total 958 patients were involved and underwent open heart surgery (MVR=728, AVR=104, DVR=54, Isolated tricuspid valve=16, MVR+TVR=56) for valvular heart disease with age ranges from 14 year to 68-year including 618 Female and 340 male (Table-1 & 2)

**Table 1.** Baseline demographic profile

| SN | Baseline Variable | value |
| --- | --- | --- |
| 1. | Age (years) | 14-68 |
| 2. | Sex |  |
|  | Male (n) | 340 |
|  | Female (n) | 618 |
| 3. | BMI (kg/m<sup>2</sup>) (mean± SD) | 14 ± 4 |

**Table 2:**
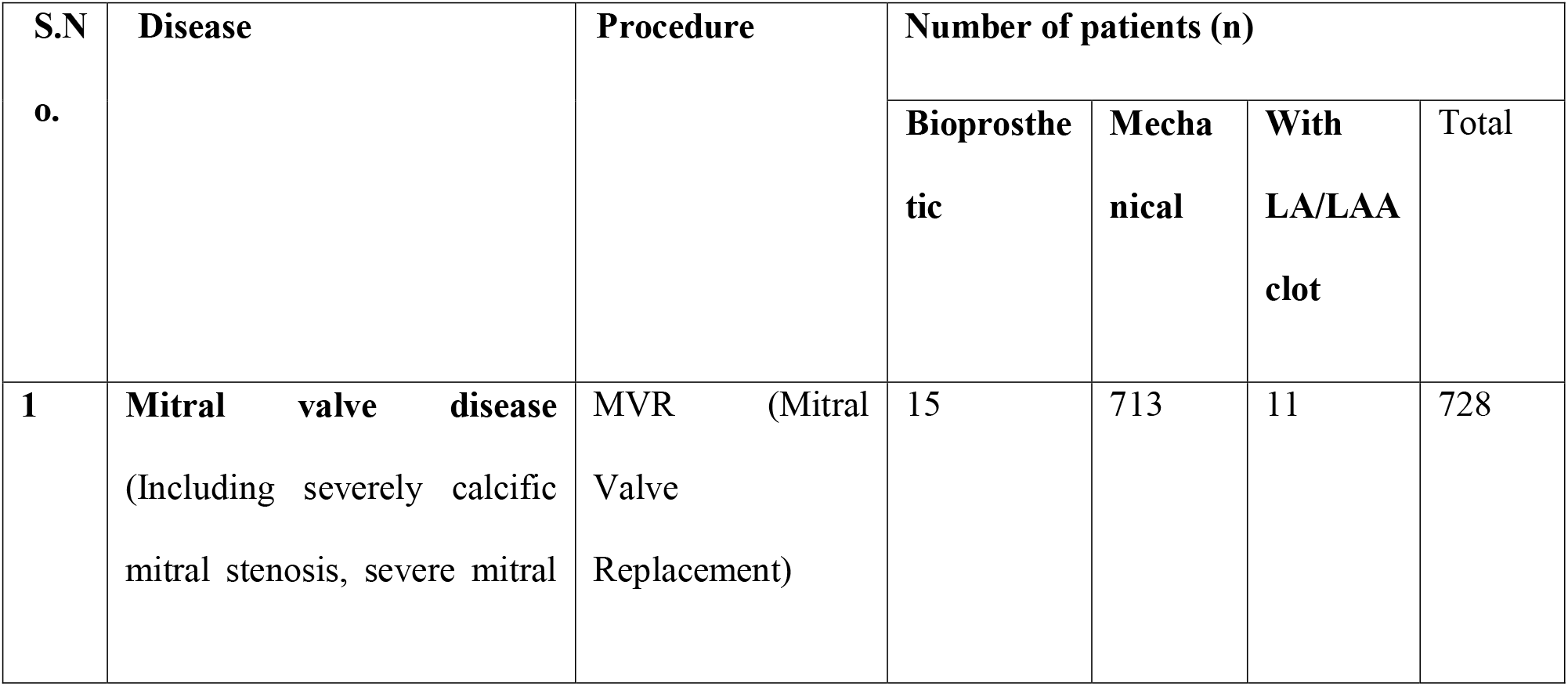
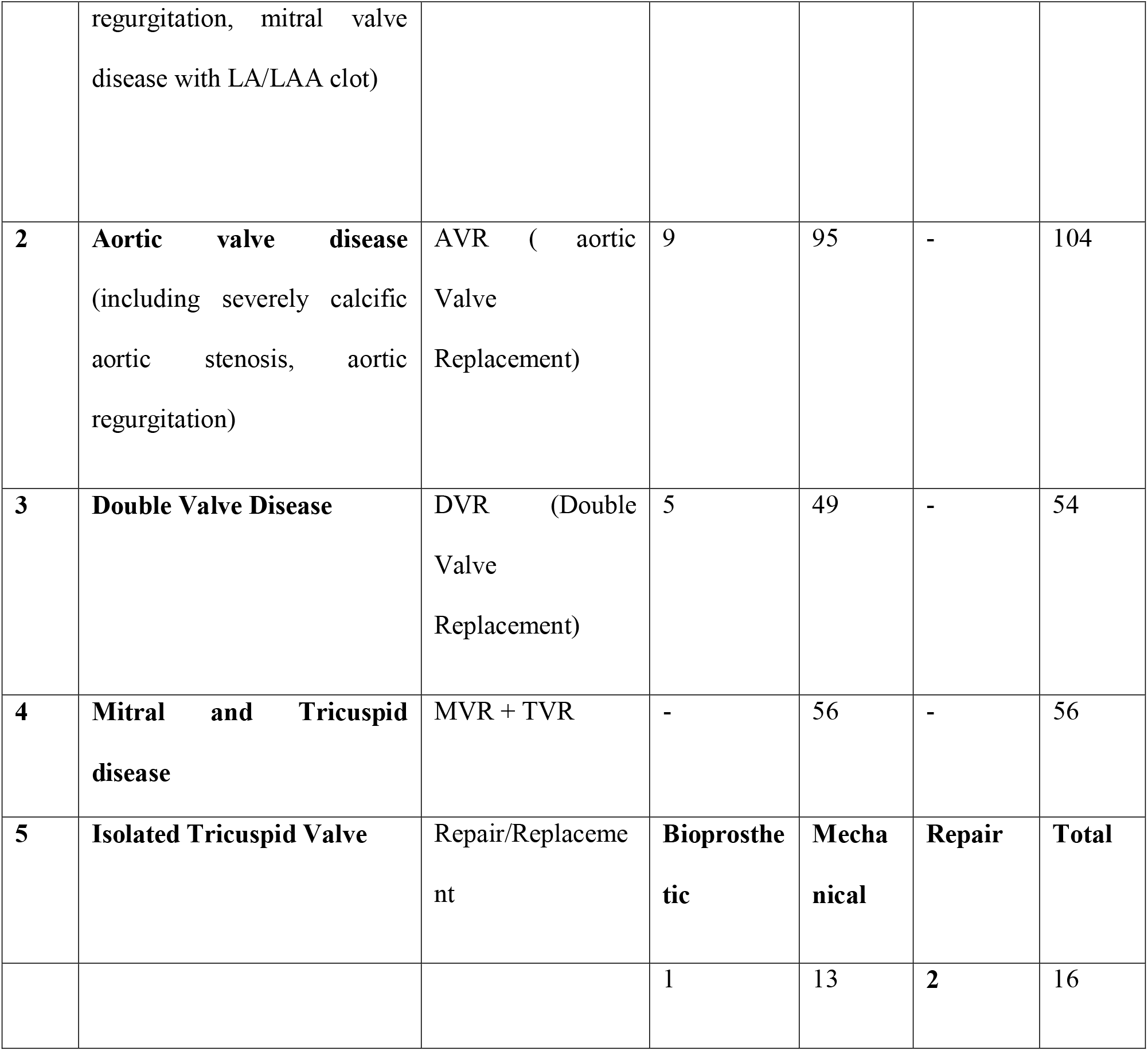
Procedures Performed Using A Small Right Anterolateral Thoracotomy Approach

| S.N<br>o. | Disease | Procedure | Number of patients (n) |  |  |  |
| --- | --- | --- | --- | --- | --- | --- |
|  |  |  | Bioprosthetic | Mechanical | With LA/LAA clot | Total |
| 1 | Mitral valve disease<br>(Including severely calcific mitral stenosis, severe mitral) | MVR (Mitral Valve Replacement) | 15 | 713 | 11 | 728 |
|  | regurgitation, mitral valve disease with LA/LAA clot) |  |  |  |  |  |
| **2** | **Aortic valve disease**<br>(including severely calcific aortic stenosis, aortic regurgitation) | AVR ( aortic Valve Replacement) | 9 | 95 | - | 104 |
| **3** | **Double Valve Disease** | DVR (Double Valve Replacement) | 5 | 49 | - | 54 |
| **4** | **Mitral and Tricuspid disease** | MVR + TVR | - | 56 | - | 56 |
| **5** | **Isolated Tricuspid Valve** | Repair/Replaceme<br>nt | **Bioprosthe**<br>**tic** | **Mecha**<br>**nical** | **Repair** | **Total** |
|  |  |  | 1 | 13 | 2 | 16 |

In our study, Mitral Valve Replacement (MVR) was administered to a cohort of 728 patients (Fig-2). Among these, 15 individuals with Bioprosthetic valves, 713 with Mechanical valves, and 11 underwent the procedure in the presence of LA/LAA clot (Fig-2). Aortic Valve Replacement (AVR) (Fig-3) was performed on 104 patients, with 9 received Bioprosthetic valves and 95 were done with Mechanical valves. Fifty four patients were operated for DVR (Fig-4) in which 5 patients were done with Bioprosthetic valves and 49 patients were done with Mechanical valves. Additionally, a combined procedure of Mitral Valve Replacement and Tricuspid Valve Replacement (MVR + TVR) was conducted on 56 patients, all of whom received Mechanical valves. For cases involving Isolated Tricuspid Valve Repair/Replacement, a total of 16 patients underwent the procedure, with one individual was done with Bioprosthetic valve, 13 received Mechanical valves, and 2 patients undergone for a Repair procedure.

**Fig 2.** a.Exposure of Mitral b. Replacement of Mitral c. LA clot

**Fig 3.** Exposure of Aortic and AVR

**Fig 4.** a. DVR b.Post operative pics

In our study there was no procedure related mortality was observed (Table-3). Smooth cannulation and satisfactory intracardiac exposure were achieved in all patients. No stroke, No renal failure, no re-exploration, no perioperative mortality, no early mortality, no conversion to median sternotomy (Table-3).

**Table 3:**
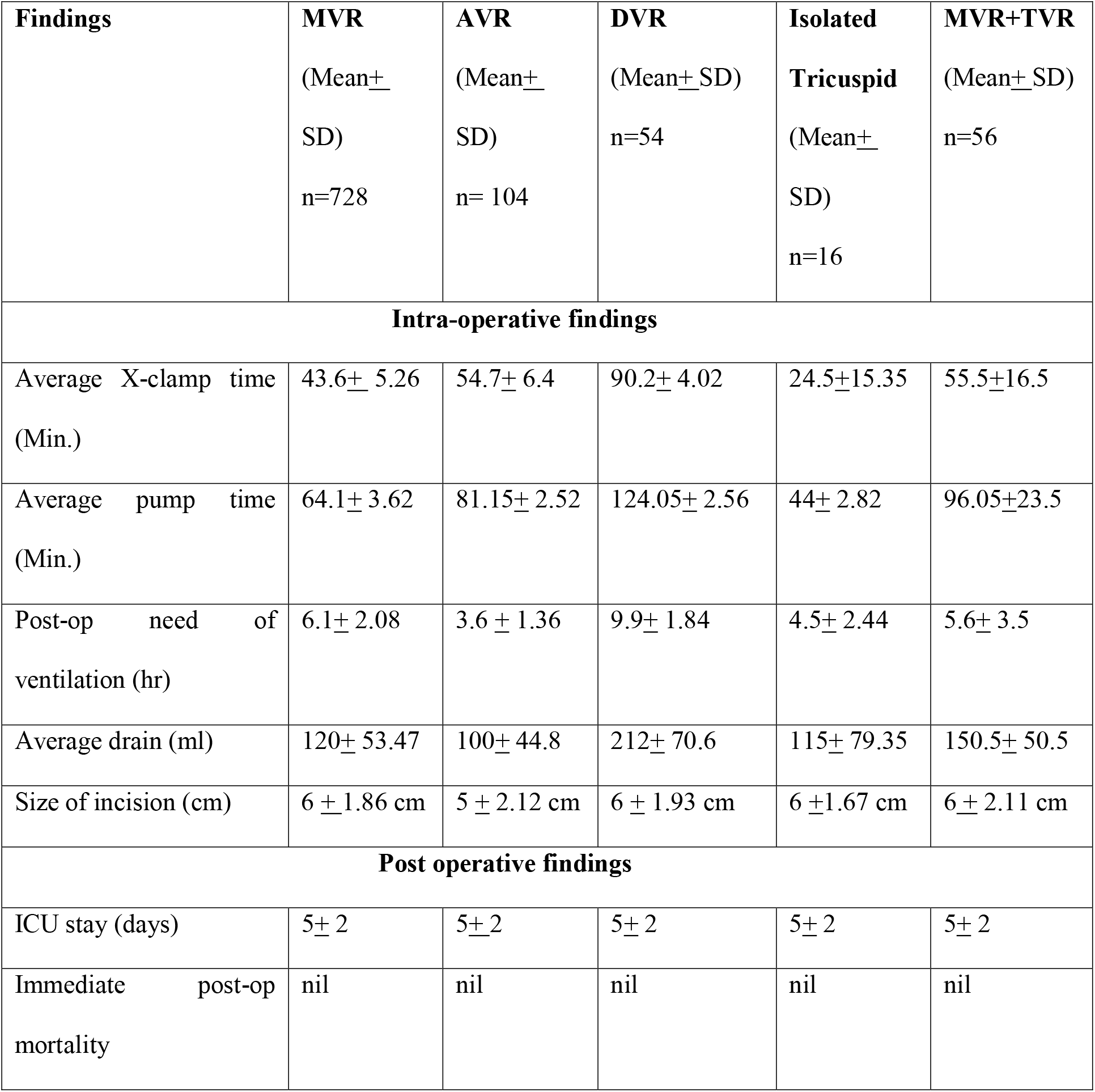
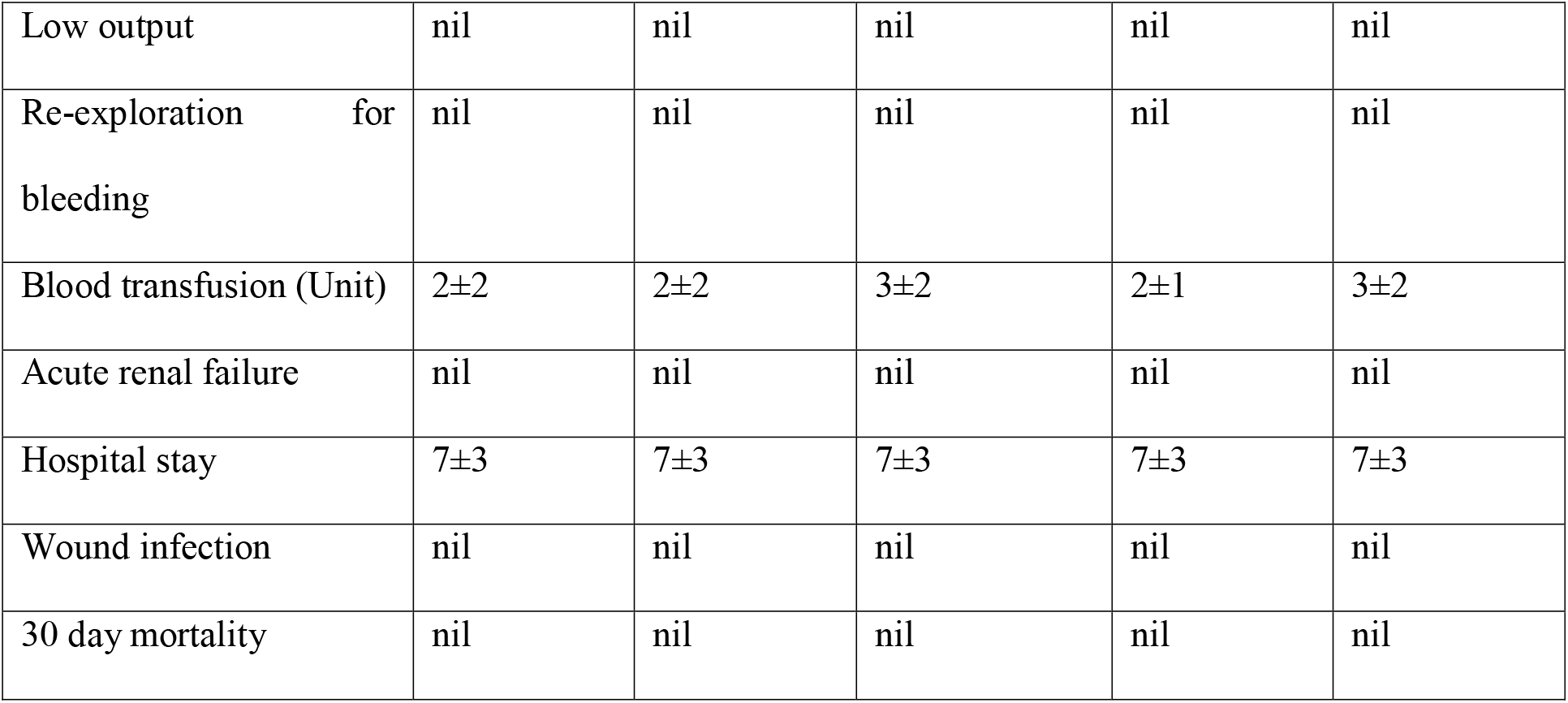
Intra operative and post-operative findings

| **Findings** | **MVR**<br>(Mean±<br>SD)<br>n=728 | **AVR**<br>(Mean±<br>SD)<br>n= 104 | **DVR**<br>(Mean±SD)<br>n=54 | **Isolated**<br>**Tricuspid**<br>(Mean±<br>SD)<br>n=16 | **MVR+TVR**<br>(Mean±SD)<br>n=56 |
| --- | --- | --- | --- | --- | --- |
| **Intra-operative findings** |  |  |  |  |  |
| Average X-clamp time<br>(Min.) | 43.6± 5.26 | 54.7± 6.4 | 90.2± 4.02 | 24.5±15.35 | 55.5±16.5 |
| Average pump time<br>(Min.) | 64.1± 3.62 | 81.15± 2.52 | 124.05± 2.56 | 44± 2.82 | 96.05±23.5 |
| Post-op need of<br>ventilation (hr) | 6.1± 2.08 | 3.6 ± 1.36 | 9.9± 1.84 | 4.5± 2.44 | 5.6± 3.5 |
| Average drain (ml) | 120± 53.47 | 100± 44.8 | 212± 70.6 | 115± 79.35 | 150.5± 50.5 |
| Size of incision (cm) | 6 ±1.86 cm | 5 ± 2.12 cm | 6 ± 1.93 cm | 6 ±1.67 cm | 6± 2.11 cm |
| **Post operative findings** |  |  |  |  |  |
| ICU stay (days) | 5± 2 | 5±2 | 5± 2 | 5± 2 | 5± 2 |
| Immediate post-op<br>mortality | nil | nil | nil | nil | nil |
| Low output | nil | nil | nil | nil | nil |
| Re-exploration for bleeding | nil | nil | nil | nil | nil |
| Blood transfusion (Unit) | 2±2 | 2±2 | 3±2 | 2±1 | 3±2 |
| Acute renal failure | nil | nil | nil | nil | nil |
| Hospital stay | 7±3 | 7±3 | 7±3 | 7±3 | 7±3 |
| Wound infection | nil | nil | nil | nil | nil |
| 30 day mortality | nil | nil | nil | nil | nil |

## DISCUSSION

Minimally-invasive cardiac valve surgery has ushered in a significant paradigm shift in recent years. Initially met with resistance from traditionalists, who argued that smaller incisions might compromise exposure and lead to inferior outcomes, this approach has evolved rapidly over the past decade. It’s now widely recognized that minimally-invasive techniques can yield results at least on par with conventional open valve surgery when performed by experienced centers. Numerous reports on minimal invasive valve procedures have surfaced over the last ten years.

One such surgical technique involves a right anterolateral thoracotomy, offering excellent valve exposure (Fig-4) and a conducive working environment while significantly reducing invasiveness and enhancing cosmetic outcomes. Patients experience less postoperative pain, shorter hospital stays, and notably reduced trauma compared to conventional incisions. Furthermore, this approach helps prevent sternal infections and often results in less blood loss both during and after the procedure. The cosmetic benefits of an incision under the right breast fold are also noteworthy. In a study by Casselman et al. [19], approximately 99% of patients reported their scars as aesthetically pleasing, which aligns with our own findings.

Over the past decade, the landscape of surgical correction for valvular heart disease has undergone significant transformation, with various surgical techniques emerging. We examined clinical data from 958 patients who underwent valve replacement via anterolateral thoracotomy with central cannulation. The advantages of the right anterolateral thoracotomy (RAT) approach are numerous, including improved cosmetic appearance, reduced risk of keloid and hypertrophic scarring, easier wound infection management, and quicker patient mobilization [6, 7]. Shorter hospital stays, early discharges, and faster returns to daily activities have a substantial positive impact on patient healthcare expenditures, which holds particular significance in our country. Notably, the psychological consequences of unaesthetic scarring, especially among young patients, are more pronounced, making our approach especially popular. Cosmetic results are of paramount importance, particularly for girls and women in India, where surgical scars can carry societal implications such as marriage taboos [9].

In light of these considerations, minimally-invasive and cosmetically favorable trans-catheter interventions have gained popularity among patients, sometimes even at the expense of slightly suboptimal results [10, 11]. Moreover, robotic-assisted minimally invasive approaches for valvular heart disease repair have also emerged [12], aiming to enhance success rates, improve cosmetic outcomes, and reduce surgical trauma. However, relevant literature on this topic remains scarce.

Notable studies such as Mishaly et al.’s [5] work in 2008, which focused on patients undergoing right anterior minithoracotomy for congenital cardiac defect repair, have provided valuable insights. While peripheral cannulation was the approach in their series, we adopted central cannulation via the thoracotomy incision. Similar efforts from other groups worldwide, such as Liu et al. from Beijing [13], have reported success in repairing more complex defects in children and adolescents. Wang et al. [4] reported positive results in patients with ventricular septal defects repaired via a minimal right vertical infra-axillary thoracotomy, with no deaths or complications from incisional wound infections or arrhythmias, which aligns with our findings.

Yamada et al. [14] compared the early postoperative quality of life between minithoracotomy and conventional sternotomy, yielding results consistent with our series. Although most of these series adopted peripheral cannulation, our technical approach involving central aortic cannulation, asymmetric breast development, defibrillation, and ventricle de-airing is well-documented. Hong et al. [15], in their 2018 publication, demonstrated that the surgical success rate for VSD repair via right sub-mammary thoracotomy and right vertical infra-axillary thoracotomy was similar to median sternotomy, suggesting these alternative methods can achieve satisfactory clinical results. Importantly, they recommended these approaches for VSD surgical repair. Our series, using the same policy of cannulation and cross clamp, yielded similar results. Gaetano et al. [7] from Italy published a series with a similar policy to ours, using anterolateral thoracotomy with central cannulation for radical correction of congenital heart disease and obtained similar results.

For female patients, Jung et al. [16] advocated the anterolateral minithoracotomy as the incision access, closely resembling our protocols. However, some reports have suggested that this incision may dissect breast tissue and lead to asymmetrical development and reduced nipple sensitivity. To mitigate these side effects, we adopted a modified incision, placing it in the sub-mammary fold (see Fig-4). Furthermore, we ensured that the incision was at least 2 to 3 cm below the mammary areola in patients with undeveloped breasts. In our anterolateral thoracotomy series, the incision ranged from 5 to 9 cm to avoid distorting growing breast tissue, particularly in prepubescent girls. This approach minimizes interference with the development of breast tissue and the pectoralis muscle, thereby preserving aesthetics (Fig. 4).

A sternal wound infection can pose significant challenges for cardiac surgeons, often leaving behind a prominent scar and unfavorable appearance. The risk of wound infections and septic complications is notably lower with thoracotomy than with median sternotomy, virtually eliminating mediastinitis [17–19]. We contend that an inconspicuous, small lower anterolateral thoracotomy scar in the breast fold is not only more cosmetically appealing but also patient-friendly, facilitating rapid recovery, shorter ICU stays, and reduced chances of mediastinitis compared to sternal incisions.

Another critical aspect of our approach is central cannulation, which mitigates complications associated with peripheral cannulation. Our method provides excellent exposure of the right atrium, caval veins, and the ascending aorta, almost on par with a median sternotomy. It allows for predictable exposure and improved cosmetics (see Fig. 1) without requiring specialized instruments. In a country like India where surgical costs are a concern for many patients, this approach offers a safe and cost-effective alternative.

In summary, our series using the anterolateral thoracotomy with central cannulation technique for valvular heart disease surgery opens up avenues for further research, particularly in resource-limited settings like India. Aspiring cardiac surgeons can benefit from the learning curve associated with this approach if it becomes a routine part of surgical units. This approach has the potential to transform cardiac surgery, offering patients enhanced cosmetic outcomes, faster recoveries, and reduced medical costs.

In 2020, Luo ZR [23] et al. conducted a comparative analysis, examining outcomes in patients undergoing ASD repair through median sternotomy, thoracotomy, and the infra-axillary approach. The findings of their study aligned with those of our research. Additionally, a separate study by Zhong Wu et al. [22] in 2012 explored mitral valve repair using an infra-axillary thoracotomy incision, demonstrating comparable excellent cosmetic and clinical outcomes to our investigation.Limitations of our study

Our study was a retrospective observational study and was limited by the number of cases and the fact that it was done in a single center. As we analyzed only collected data of operated patients and we did not compare with any control group, absence of control group is limitation of our study. Prospective randomized controlled studies with a larger sample size, even multicenter cooperation, must be conducted to confirm the results. In addition, a longer follow-up is essential, especially for those patients with thoracic deformity.

## Conclusions

All the data showed that conventional aorto-caval cannulation from antero-lateral thoracotomy approach for minimal invasive open-heart surgery for valvular heart disease is also a good procedure to minimize the side effects of sternotomy and femoral cannulation. This procedure is safe to provide the same quality of treatment through a less traumatic and better cosmetic incision resulting in less hospital stay and a lower overall cost. This is likely the world’s largest series of valvular heart surgeries conducted via a thoracotomy approach with central cannulation, concluding that this procedure is safe and reproducible.

## Data Availability

All data produced in the present study are available upon reasonable request to the authors

## Acknowledgements

**Ethical approval was taken by ethical committee of institute**

